# Most users of a prenatal consultation dedicated to future fathers are immigrants facing hardship

**DOI:** 10.1101/2023.09.28.23295896

**Authors:** Pauline Penot, Gaëlle Jacob, Audrey Guerizec, Clotilde Trevisson, Valérie-Anne Letembet, Raya Harich, Thomas Phuong, Bruno Renevier, Pierre-Etienne Manuellan, Annabel Desgrées du Loû, the Partage Study Group

## Abstract

**Background:** Prenatal care provides pregnant women with repeated opportunities for prevention, screening and diagnosis that have no current extension to future fathers. It also contributes to women’s general better access to health. The goal of PARTAGE study was to evaluate the level and determinants of adherence to a prenatal prevention consultation dedicated to men.

**Methods:** Between January 2021 and April 2022, we conducted a monocentric interventional study in Montreuil hospital. We assessed the acceptance of a prenatal prevention consultation newly offered to every future father, through their pregnant partner’s prior consent to provide their contact details.

**Results:** 3,038 women provided contact information used to reach the fathers; effective contact was established with 2,516 men, of whom 1,333 (53%) came for prenatal prevention consultation. Immigrant men were more likely to come than French-born men (56% versus 49%, p < 0·001), and the more they faced social hardship, the more likely they were to accept the offer. In multivariate analysis, men born in Subsaharan Africa and Asia were twice as likely to attend the consultation as those born in Europe or North America.

**Conclusion:** Acceptance of this new offer was high. Moreover, this consultation was perceived by vulnerable immigrant men as an opportunity to integrate a healthcare system they would otherwise remain deprived of.

**Trial Registration:** https://classic.clinicaltrials.gov/ct2/show/NCT05085717

## Introduction

### Gynecological care: an opportunity for prevention, screening and health improvement

Worldwide, men have less contact with the healthcare system (1) and seek primary care less offen than women (2) (3) (4). One reason is that gynecological care (contraception, pregnancy, cervical cancer prevention) makes women used to meeting healthcare professionals regularly, even in absence of any disease. (5).

Most countries recommend regular medical check-ups and biological monitoring during pregnancy. When the father’s health is taken into the picture, this is to avoid genetical or infectious transmission rather than keeping the father himself in good health (6). Even then, infectious diseases are poorly screened in the second parent : HIV diagnosis is delayed in heterosexual men compared to women, because male partner uptake of HIV testing during antenatal care remains scarce (7), despite numerous initiatives to promote it (8) (9) (10) (11) (12).

Prenatal visits are also used to encourage behavioral changes in pregnant women, such as smoking and alcoholic withdrawal, promoting physical exercise and combating overweight (13) (14). To date, men do not have such opportunities. Yet, involving their partner supports behavioral changes in pregnant women (15). In addition, important health needs were identified in a cohort of future fathers (16) and fatherhood is a high-risk period for depression in men (17) (18).

Expecting a child could therefore be an opportunity to offer men a consultation designed to protect the mother and the foetus from transmissible diseases, but also to improve or preserve their own health (19).

### The French context and the Seine Saint Denis specific situation

In France, seven medical visits are scheduled during a pregnancy. The prenatal HIV screening strategy is highly effective in women, with at least 97% of them tested at every pregnancy (20). Systematic HIV screening is also recommended for fathers (21), yet not performed: between 2017 and 2020, among 567 fathers surveyed, only 3% knew a man who had been tested for HIV in connection with his partner’s pregnancy (22).

The French health insurance scheme mentions the possibility of a medical examination for the father. However, there are no guidelines for reimbursing this consultation or the biological tests that would be prescribed during it: paternal prenatal consultation is therefore not currently practiced.

Seine Saint-Denis (population; 1.6 million inhabitants), located in the northeastern suburbs of Paris (Ile de France region), is the department with the highest rates of immigration and poverty in mainland France (23). Seine Saint Denis’ tertiary care maternity unit is located in Montreuil: it provides prenatal care for some 3,500 women every year. In 2020-2021, 4·6% of women who gave birth at Montreuil hospital were immigrants with < 12 months’ residence in France (24). 52% of women who delivered their child in Montreuil hospital over the same period were later identified as socially vulnerable, 41% of them being born in Subsaharan Africa and 11% in Asia (25).

In this context where poverty and immigration prevail, future fathers hardly get targetted by interventions aimed at reducing social inequalities in health. We consequently built an interventional research to assess the take-up rate and the factors associated with acceptance of a prenatal consultation dedicated to them.

## Methods

PARTAGE (« Prévention, Accès aux droits, Rattrapage vaccinal, Traitement des Affections pendant la Grossesse et pour l’Enfant ») was a monocentric interventional research without control arm assesssing for 15 months (January 2021-April 2022) the acceptability of a new prenatal prevention consultation offered through the pregnant women to all future fathers in Montreuil’s hospital.

### Study population

Eligible women were over 18 pregnant women attending a first prenatal consultation at Montreuil hospital during the study period, reporting a male partner positively involved in the pregnancy.

Eligible men were those designated by the participating woman as the father, provided they lived in Ile-de-France region. To make it simple, « men », « partners » or « fathers » all refer to them.

### Intervention

The interview was integrated into the women’s maternity trajectory. Trained interviewers offered eligible women to respond to a face-to-face questionnaire that included their sociodemographic and marital characteristics (maternal questionnaire, MQ, additional file 1). Women were asked to provide the future father’s contact details. When the father was present, the project was presented directly to the couple and the consultation appointment could be made immediately.

An information letter was given to all eligible pregnant women. An e-mail was sent to men for whom an e-mail address was provided, with an online appointment plateform link and dedicated phone-line and e-mail address to schedule consultation. Fathers who did not reply were called by a research midwife and offered an appointment either during the day or in the evening, on weekdays or saturday, at the hospital or in the city center, within easy reach of public transportation.

Fathers received a text reminder before appointment. If they failed to attend, they were systematically called again and given the option of either refusing the consultation (their reason for refusal was then collected and they were not called again) or rescheduling and being called again in case of another missed appointment.

Fathers were seen by a doctor or midwife. Blood pressure was measured, clinical examination was carried out if needed, biological tests were adapted to individual’s exposure and medical history. Vaccines were updated; fathers lacking health insurance coverage met a social worker to obtain it. Test results were delivered face to face, by telephone or by e-mail, according to men’s choice. Depending on their needs, men were referred to a health mediator, a psychologist, a general practioner or any hospital specialist.

The full protocol is available on ClinicalTrials.gov: https://classic.clinicaltrials.gov/ct2/show/NCT05085717.

### Ethics

The French Data Protection Authority (CNIL, registration number 921135) and the French Personnal Data Protection Comittee (Comite de Protection des Personnes, registation number 21.01.19.44753 amended by 21.05022.944753 decision, allowing and increased number of inclusions and additional data collection) gave full ethical approval.

### Statistical analysis

Women’s own sociodemographic characteristics, their partners’ ones as well as data on the couple (lenght of relationship, previous children, type of union, communication about sexually transmitted infections and previous screening) were collected in maternal questionnaire. In addition to the general sociodemographic data; variables exclusively or particularly related to immigrant status were collected (main reason for women’s migration, lenght of stay, permit of residence, housing conditions, health insurance coverage).

Data were compared between fathers who attended the prenatal prevention consultation and those who did not, using Chi [2] or Fisher exact tests for categorical variables and Student’s or Wilcoxon rank sum tests for quantitative variables, as appropriate. All tests were two-sided with p-values < 0·05 defined as statistically significant. Multivariate logistic regression models were built to identify the factors associated with men’s take-up of this offer, based on their own characteristics on one hand, and on their pregnant partner’s as well as the couple’s characteristics on the other hand, in the general study population, and then in the sub-groups of immigrant men and women. Backward elimination procedures were used to determine the final models. Given the limited number of missing data, no imputation was performed, in order to avoid introducing bias: incomplete observations were removed from the multivariate models. Analyses were performed in Stata SE 17 (Stata Corporation, College Station, TX, USA).

## Results

### 1. Acceptance rate and characteristics of the studied population

Among 4205 women with first hospital prenatal visit during the study period, 3808 were eligible, 3038 provided effective contact details wich enabled the research team to contact their partner. Contact was actually made (face-to-face, by phone or via message exchange) with 2516 men (83%), of whom 1333 (53%) eventually attended father’s prenatal prevention consultation (FPPC) (Figure 1).

**Figure 1:**
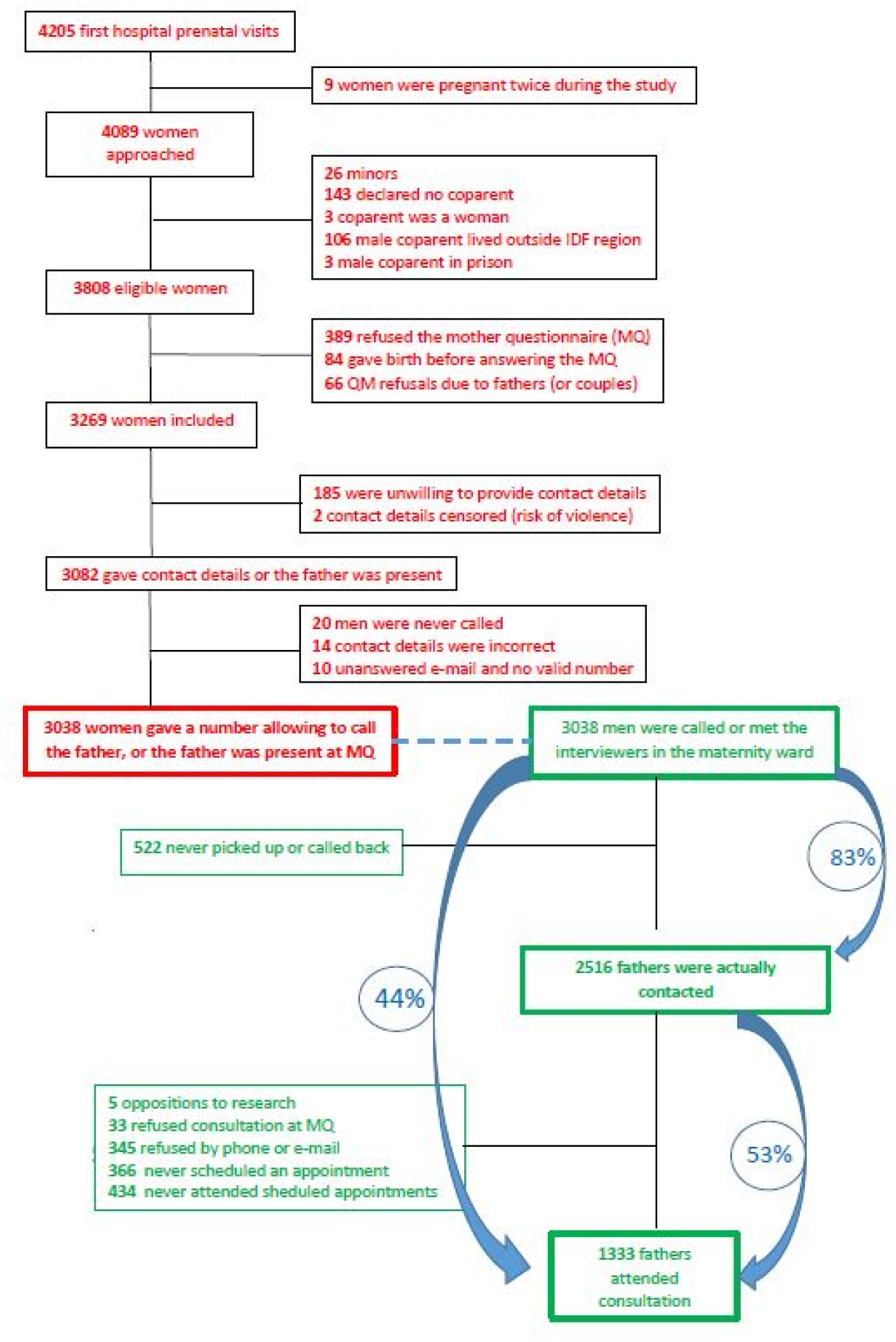
Study flow chart, Montreuil (France) 2021-2022.

**Regarding women:** their median age was 31 years, interquartile range [] 28-35 years. One third was expecting a first child; 49% had a university degree, whereas 46% were unemployed prior to current pregnancy; 13% believed they had never been tested for HIV, while 15% identified their ongoing pregnancy serology as their first. Only 36% recalled a couple’s discussion about HIV or sexually transmitted infections (STIs), and 56% didn’t know if their partner had ever been tested for HIV. 78% predicted that the future father would attend the prenatal consultation he would be offered.

Fifty-two percent were immigrants, most often from Subsaharan Africa (20%), North Africa or the Middle East (18%). Among the immigrant women (N = 1590): 51% had been in France for less than 7 years; 19% had no residence permit and 9% no health insurance coverage. Two-thirds had come to France to join either their husbands (43%) or a family member (26%), while 17% had come to seek a better life, 8% to study, 6% to escape threats or violence, and only 0.7% for health reasons (table 1).

**Regarding men:** Their median age was 35 [30-40]; 35% had superior education, 14% were unemployed. Fifty-nine percent of those with whom contact had been established were immigrants (N= 1490): 25% came from Subsaharan Africa, 19% from North Africa or the Middle East. Only 29% had been in France for less than 7 years; 18% had no residence permit, and 11% had no health insurance coverage.

**Table 1:**
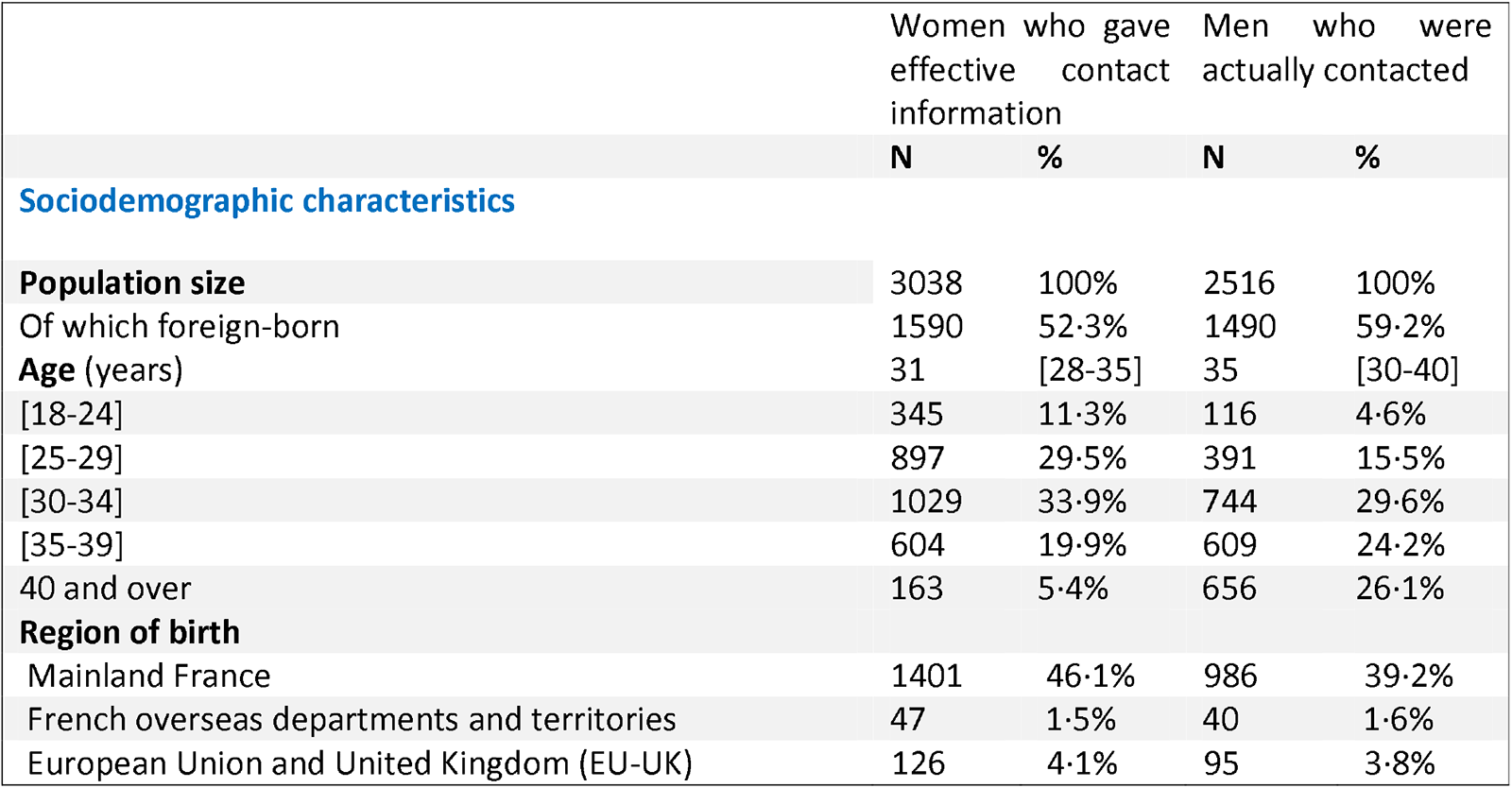
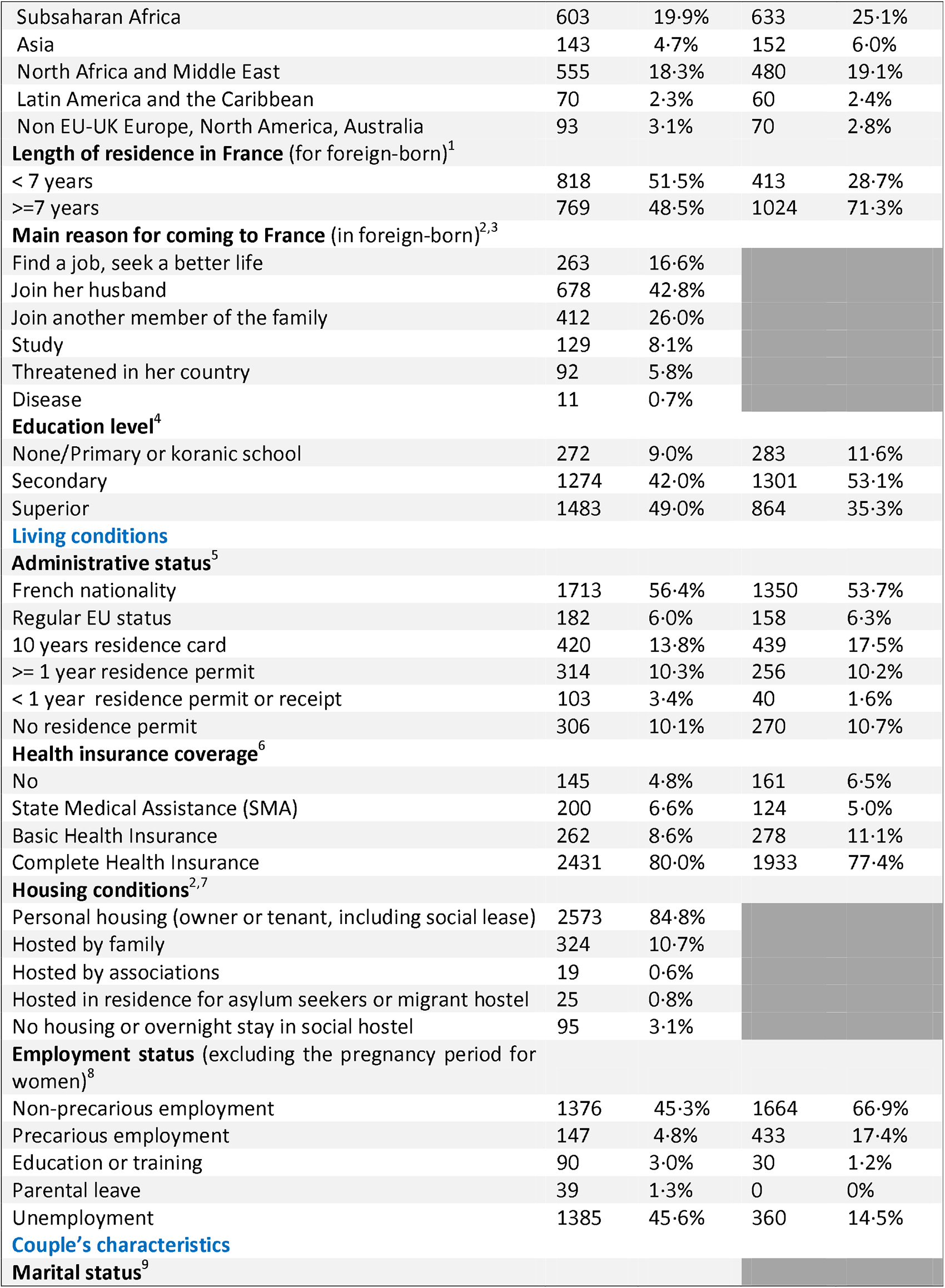
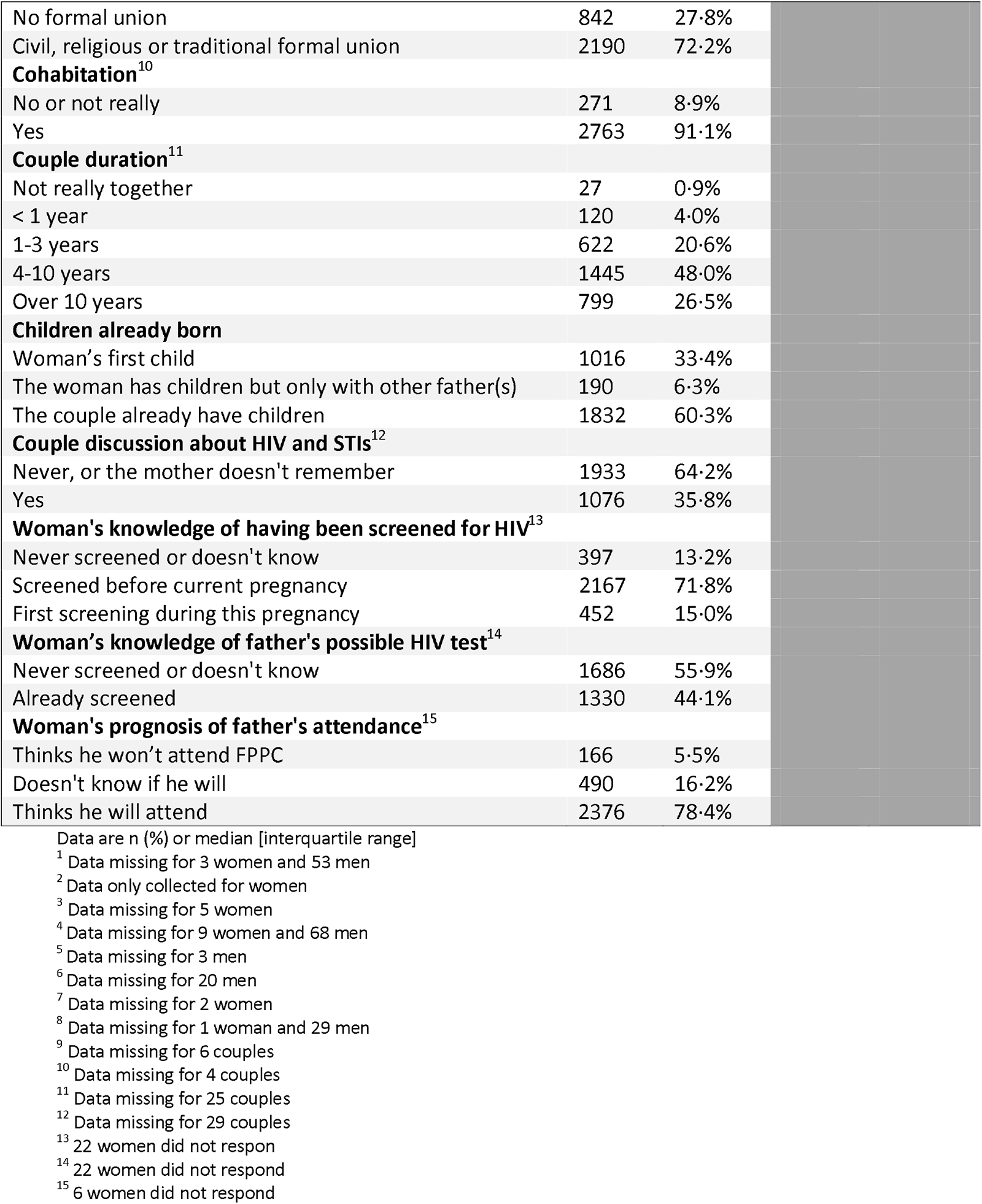
Characteristics of the study population, Montreuil (France) 2021-2022.

|  | Women who gave effective contact information |  | Men who were actually contacted |  |
| --- | --- | --- | --- | --- |
|  | N | % | N | % |
| **Sociodemographic characteristics** |  |  |  |  |
| **Population size** | 3038 | 100% | 2516 | 100% |
| Of which foreign-born | 1590 | 52.3% | 1490 | 59.2% |
| **Age (years)** | 31 | [28-35] | 35 | [30-40] |
| [18-24] | 345 | 11.3% | 116 | 4.6% |
| [25-29] | 897 | 29.5% | 391 | 15.5% |
| [30-34] | 1029 | 33.9% | 744 | 29.6% |
| [35-39] | 604 | 19.9% | 609 | 24.2% |
| 40 and over | 163 | 5.4% | 656 | 26.1% |
| **Region of birth** |  |  |  |  |
| Mainland France | 1401 | 46.1% | 986 | 39.2% |
| French overseas departments and territories | 47 | 1.5% | 40 | 1.6% |
| European Union and United Kingdom (EU-UK) | 126 | 4.1% | 95 | 3.8% |
| Subsaharan Africa | 603 | 19.9% | 633 | 25.1% |
| Asia | 143 | 4.7% | 152 | 6.0% |
| North Africa and Middle East | 555 | 18.3% | 480 | 19.1% |
| Latin America and the Caribbean | 70 | 2.3% | 60 | 2.4% |
| Non EU-UK Europe, North America, Australia | 93 | 3.1% | 70 | 2.8% |
| **Length of residence in France (for foreign-born)<sup>1</sup>** |  |  |  |  |
| < 7 years | 818 | 51.5% | 413 | 28.7% |
| >=7 years | 769 | 48.5% | 1024 | 71.3% |
| **Main reason for coming to France (in foreign-born)<sup>2,3</sup>** |  |  |  |  |
| Find a job, seek a better life | 263 | 16.6% |  |  |
| Join her husband | 678 | 42.8% |  |  |
| Join another member of the family | 412 | 26.0% |  |  |
| Study | 129 | 8.1% |  |  |
| Threatened in her country | 92 | 5.8% |  |  |
| Disease | 11 | 0.7% |  |  |
| **Education level<sup>4</sup>** |  |  |  |  |
| None/Primary or koranic school | 272 | 9.0% | 283 | 11.6% |
| Secondary | 1274 | 42.0% | 1301 | 53.1% |
| Superior | 1483 | 49.0% | 864 | 35.3% |
| **Living conditions** |  |  |  |  |
| **Administrative status<sup>5</sup>** |  |  |  |  |
| French nationality | 1713 | 56.4% | 1350 | 53.7% |
| Regular EU status | 182 | 6.0% | 158 | 6.3% |
| 10 years residence card | 420 | 13.8% | 439 | 17.5% |
| >= 1 year residence permit | 314 | 10.3% | 256 | 10.2% |
| < 1 year residence permit or receipt | 103 | 3.4% | 40 | 1.6% |
| No residence permit | 306 | 10.1% | 270 | 10.7% |
| **Health insurance coverage<sup>6</sup>** |  |  |  |  |
| No | 145 | 4.8% | 161 | 6.5% |
| State Medical Assistance (SMA) | 200 | 6.6% | 124 | 5.0% |
| Basic Health Insurance | 262 | 8.6% | 278 | 11.1% |
| Complete Health Insurance | 2431 | 80.0% | 1933 | 77.4% |
| **Housing conditions<sup>2,7</sup>** |  |  |  |  |
| Personal housing (owner or tenant, including social lease) | 2573 | 84.8% |  |  |
| Hosted by family | 324 | 10.7% |  |  |
| Hosted by associations | 19 | 0.6% |  |  |
| Hosted in residence for asylum seekers or migrant hostel | 25 | 0.8% |  |  |
| No housing or overnight stay in social hostel | 95 | 3.1% |  |  |
| **Employment status (excluding the pregnancy period for women)<sup>8</sup>** |  |  |  |  |
| Non-precarious employment | 1376 | 45.3% | 1664 | 66.9% |
| Precarious employment | 147 | 4.8% | 433 | 17.4% |
| Education or training | 90 | 3.0% | 30 | 1.2% |
| Parental leave | 39 | 1.3% | 0 | 0% |
| Unemployment | 1385 | 45.6% | 360 | 14.5% |
| **Couple's characteristics** |  |  |  |  |
| **Marital status<sup>9</sup>** |  |  |  |  |

|  |  |  |
| --- | --- | --- |
| No formal union | 842 | 27·8% |
| Civil, religious or traditional formal union | 2190 | 72·2% |
| **Cohabitation<sup>10</sup>** |  |  |
| No or not really | 271 | 8·9% |
| Yes | 2763 | 91·1% |
| **Couple duration<sup>11</sup>** |  |  |
| Not really together | 27 | 0·9% |
| < 1 year | 120 | 4·0% |
| 1-3 years | 622 | 20·6% |
| 4-10 years | 1445 | 48·0% |
| Over 10 years | 799 | 26·5% |
| **Children already born** |  |  |
| Woman's first child | 1016 | 33·4% |
| The woman has children but only with other father(s) | 190 | 6·3% |
| The couple already have children | 1832 | 60·3% |
| **Couple discussion about HIV and STIs<sup>12</sup>** |  |  |
| Never, or the mother doesn't remember | 1933 | 64·2% |
| Yes | 1076 | 35·8% |
| **Woman's knowledge of having been screened for HIV<sup>13</sup>** |  |  |
| Never screened or doesn't know | 397 | 13·2% |
| Screened before current pregnancy | 2167 | 71·8% |
| First screening during this pregnancy | 452 | 15·0% |
| **Woman's knowledge of father's possible HIV test<sup>14</sup>** |  |  |
| Never screened or doesn't know | 1686 | 55·9% |
| Already screened | 1330 | 44·1% |
| **Woman's prognosis of father's attendance<sup>15</sup>** |  |  |
| Thinks he won't attend FPPC | 166 | 5·5% |
| Doesn't know if he will | 490 | 16·2% |
| Thinks he will attend | 2376 | 78·4% |

Data are n (%) or median [interquartile range]
<sup>1</sup> Data missing for 3 women and 53 men
<sup>2</sup> Data only collected for women
<sup>3</sup> Data missing for 5 women
<sup>4</sup> Data missing for 9 women and 68 men
<sup>5</sup> Data missing for 3 men
<sup>6</sup> Data missing for 20 men
<sup>7</sup> Data missing for 2 women
<sup>8</sup> Data missing for 1 woman and 29 men
<sup>9</sup> Data missing for 6 couples
<sup>10</sup> Data missing for 4 couples
<sup>11</sup> Data missing for 25 couples
<sup>12</sup> Data missing for 29 couples
<sup>13</sup> 22 women did not respond

### 2. Characteristics of women whose partner accepted the offer

#### Comparison of women whose partner came to the consultation with women whose partner did not

Partners of primipara were more likely to come than those of women who were already mothers, especially if the couple already had children together (53% versus 38%).

Partners of women born in Subsaharan Africa, Asia and Latin America-Haiti took up the offer better than those born in France (56%, 62% and 50%, respectively, versus 38%).

Partners of women in precarious situations (homeless, with SMA or no health insurance coverage) came more often than those of women living in their own homes or registered with the general health insurance scheme.

Partners of women declaring a first HIV serology during the current pregnancy were more likely to take up the offer than those of women who were unaware of having been tested, and those who had been tested prior to the current pregnancy (50% versus 46% and 42%, respectively).

Half of men whose partners predicted they would take up the offer actually came, versus 22% of those whose partners were uncertain, and 13% of those whose partners predicted that refusal (*p < 0.001*, table 2).

**Table 2:** Women’s characteristics by partner’s attendance to father prenatal prevention consultation, Montreuil (France) 2021-2022 (N=3038 partners called or met)

|  | Women whose partner came (N=1333) |  | Women whose partner did not come (N = 1705) |  | P-value |
| --- | --- | --- | --- | --- | --- |
|  | n | % | n | % |  |
| **Age<sup>1</sup>** |  |  |  |  | 0.531 |
| [18-24 years] | 153 | 44.3% | 192 | 55.7% |  |
| [25-29 years] | 376 | 41.9% | 521 | 58.1% |  |
| [30-34 years] | 456 | 44.3% | 573 | 55.7% |  |
| 35 and over | 348 | 45.4 % | 419 | 54.6% |  |

| Region of birth |  |  |  |  |  |
| --- | --- | --- | --- | --- | --- |
| France | 557 | 38.5% | 891 | 61.5% | <0.001 |
| UE 27-UK | 54 | 42.9% | 72 | 57.1% |  |
| Europe non-EU 27-UK, North America, Australia | 39 | 41.9% | 54 | 58.1% |  |
| North Africa and Middle East | 221 | 39.8% | 334 | 60.2% |  |
| Subsaharan Africa | 338 | 56.0% | 265 | 44.0% |  |
| Asia | 89 | 62.2% | 54 | 37.8% |  |
| Latin America and the Caribbean | 35 | 50.0% | 35 | 50.0% |  |
| Length of residence in France<sup>2</sup> |  |  |  |  |  |
| < 7 years | 443 | 54.2% | 374 | 45.8% | <0.001 |
| >= 7 years | 331 | 43.1% | 437 | 56.9% |  |
| Born in France | 557 | 38.5% | 891 | 61.5% |  |
| Main reason for migration |  |  |  |  |  |
| Not applicable (born in France) | 557 | 38.5% | 890 | 61.5% | <0.001 |
| Find a job, seek a better life | 150 | 57.0% | 113 | 43.0% |  |
| Join someone (family, husband) or study | 553 | 45.3% | 667 | 54.7% |  |
| Disease or threatened in her country | 71 | 68.9% | 32 | 31.1% |  |
| Education level |  |  |  |  |  |
| None/ Primary or koranic school | 153 | 56.2% | 119 | 43.8% | <0.001 |
| Secondary | 497 | 39.0% | 777 | 61.0% |  |
| Superior | 679 | 45.8% | 804 | 54.2% |  |
| Administrative status |  |  |  |  |  |
| French nationality | 674 | 39.3% | 1039 | 60.7% | <0.001 |
| Regular EU status | 83 | 45.6% | 99 | 54.4% |  |
| 10-year residence card | 185 | 44.0% | 235 | 56.0% |  |
| Residence permit 1 year or more | 156 | 49.7% | 158 | 30.3% |  |
| < 1 year residence permit or receipt | 52 | 50.5% | 51 | 49.5% |  |
| No residence permit | 183 | 59.8% | 123 | 40.2% |  |
| Health insurance coverage |  |  |  |  |  |
| No | 79 | 54.5% | 66 | 45.5% | <0.001 |
| State Medical Assistance | 116 | 58.0% | 84 | 42.0% |  |
| Basic Health Insurance only | 96 | 36.6% | 166 | 63.4% |  |
| Complete Health Insurance | 1042 | 42.9% | 1389 | 57.1% |  |
| Employment status (excluding the pregnancy period) |  |  |  |  |  |
| Precarious employment, student, in training | 123 | 51.2% | 117 | 48.8% | 0.056 |
| Non-precarious employment | 598 | 43.5% | 778 | 56.5% |  |
| Unemployment or on parental leave | 612 | 43.1% | 809 | 56.9% |  |
| Housing conditions |  |  |  |  |  |
| Personal housing | 1090 | 42.4% | 1483 | 57.6% | <0.001 |
| Hosted (family, association) | 173 | 47.0% | 195 | 53.0% |  |
| On the street or at the 115 | 70 | 73.7% | 25 | 26.3% |  |
| Couple duration |  |  |  |  |  |
| Not really together | 10 | 37.0% | 17 | 63.0% | <0.001 |
| < 1 year | 67 | 55.8% | 53 | 44.2% |  |
| 1-3 years | 306 | 49.2% | 316 | 50.8% |  |
| 4-10 years | 638 | 44.1% | 807 | 55.9% |  |
| Over 10 years | 312 | 39.0% | 487 | 91.0% |  |
| Children already born |  |  |  |  |  |
| No | 536 | 52.8% | 480 | 47.2% | <0.001 |
| Yes, including children with the same father | 703 | 38.4% | 1129 | 61.6% |  |
| Yes, but only with other father(s) | 94 | 49.5% | 96 | 50.5% |  |
| **Have you had an HIV test?** |  |  |  |  | 0.004 |
| No or I don't think so | 182 | 45.8% | 215 | 54.2% |  |
| Yes, even before the current pregnancy | 914 | 42.2% | 1253 | 57.8% |  |
| Yes, only during this pregnancy | 228 | 50.4% | 224 | 49.6% |  |
| **Have you ever talked about HIV/STIs with your partner?** |  |  |  |  |  |
| No or I don't know | 858 | 44.4% | 1075 | 55.6% | 0.472 |
| Yes | 463 | 43.0% | 613 | 57.0% |  |
| **Do you think he will accept FPPC?** |  |  |  |  | <0.001 |
| No | 22 | 13.2% | 144 | 86.8% |  |
| I don't know | 108 | 22.0% | 382 | 78.0% |  |
| Yes | 1200 | 50.5% | 1176 | 49.5% |  |

<sup>1</sup> Cohabitation with the father, marital status, women's knowledge of father's HIV status and of any HIV performed by the father were also analyzed, with no difference between groups. A complete comparison of women's characteristics is provided as additional file 2.
<sup>2</sup> When the sum of the category headcounts does not correspond to the total headcount, there are missing data (see table 1).

#### Women’s characteristics associated with their partners’ attendance

After adjusting for other women’s characteristics, those born in Subsaharan Africa (OR 2.23 [1.79-2.78]), Asia (OR 3.14 [2.15-4.59]) or Latin America-Haiti (OR 1.71 [1.04-2.82]) were more likely to have their partners attending than those born in France. Women’s level of education remained associated with their partners’ attendance: partners of women with a low level of formal education (no or only elementary school) or, on the contrary, a high level of formal education (university degree) came more often than those of women with a high school level (figure 2, graph a).

#### Characteristics of the immigrant women sub-population associated with their partner’s attendance

In multivariate analysis, partners of women born Subsaharan Africa and Asia made greater use of this new consultation than those of women from Europe, USA or Australia, even after adjustment for length of stay, reason for migration, housing, administrative status and parenthood (OR 1.61 [1.16-2.23] and OR 2.35 [1.50-3.68], respectively). Homelessness, migration for their own safety or to seek a better life, and a residence period of less than seven years were independently associated with the partner’s attendance (figure 2, graph c).

### 3. Characteristics of men who took up the offer

#### Comparison of men who came to the consultation versus those who did not

Participation was higher among men aged 30 and over than among younger ones, and among Subsaharan African, Asian and Latin American-Haitian immigrants than among French-born men (49% versus 61%, 63% and 53% respectively). Immigrants present for less than 7 years came more often than those who had been in France for longer (66% versus 55%, p < 0.001). As for women, men with social vulnerabilities (no residence permit, no affiliation to the general health insurance scheme) made greater use of consultations than less precarious ones. The take-up rate was lower among men with a secondary level of education than among men with a lower or higher level of education (49% versus 63% and 59%, respectively, p < 0.001), and a similar trend was observed with regard to women’s level of education. Finally, the proportion of adherence was similar whether or not men had any medical follow-up (53% versus 47%, p = 609, table 3).

**Table 3:** Men’s characteristics by attendance to father prenatal prevention consultation, Montreuil (France) 2021-2022 (N=2516 men actually contacted)

|  | Men who attended<br>(N = 1333) |  | Men who did not attend despite<br>effective contact<br>(N = 1183) |  | *P-value* |
| --- | --- | --- | --- | --- | --- |
| Age<sup>1</sup> |  |  |  |  |  |
| [18-29 years] | 238 | 46.9% | 269 | 53.1% | 0.009 |
| [30-34 years] | 402 | 54.0% | 342 | 46.0% |  |
| 35 and over | 693 | 54.8% | 571 | 45.2% |  |
| Region of birth |  |  |  |  |  |
| France | 500 | 48.7% | 526 | 51.3% | <0.001 |
| EU 27 | 42 | 44.2% | 53 | 55.6% |  |
| Non-EU 27 Europe, Canada | 32 | 45.7% | 38 | 54.3% |  |
| North Africa and Middle East | 245 | 51.0% | 235 | 49.0% |  |
| Subsaharan Africa | 386 | 61.0% | 247 | 39.0% |  |
| Asia | 96 | 63.2% | 56 | 36.8% |  |
| Latin America and the Caribbean | 32 | 53.3% | 28 | 46.7% |  |
| Length of residence in France |  |  |  |  |  |
| < 7 years | 274 | 66.3% | 139 | 33.7% | <0.001 |
| >= 7 years | 559 | 54.6% | 465 | 45.4% |  |
| Born in France | 500 | 48.7% | 526 | 51.3% |  |
| Administrative status |  |  |  |  |  |
| French nationality | 679 | 50.3% | 671 | 49.7% | <0.001 |
| Regular EU status | 75 | 47.5% | 83 | 52.5% |  |
| 10-year residence card | 217 | 49.4% | 222 | 50.6% |  |
| >= 1 year residence permit | 143 | 55.9% | 113 | 44.1% |  |
| Residence permit < 1 year or receipt | 23 | 57.5% | 17 | 42.5% |  |
| No residence permit | 196 | 72.6% | 74 | 27.4% |  |
| Health insurance coverage |  |  |  |  |  |
| No | 106 | 65.8% | 55 | 34.2% | <0.001 |
| State Medical Assistance (SMA) | 88 | 71.0% | 36 | 29.0% |  |
| Basic Health Insurance | 118 | 42.5% | 160 | 57.5% |  |
| Complete Health Insurance | 1021 | 52.8% | 912 | 47.2% |  |
| Employment status<sup>5</sup> |  |  |  |  |  |
| Precarious employment, student, in training | 247 | 53.4% | 216 | 46.6% | 0.052 |
| Non-precarious employment | 872 | 52.4% | 792 | 47.6% |  |
| Unemployment | 214 | 59.4% | 146 | 40.6% |  |
| Level of education |  |  |  |  |  |
| None, primary or koranic school | 179 | 63.3% | 104 | 36.7% | <0.001 |
| Secondary | 641 | 49.3% | 660 | 50.7% |  |
| Superior | 512 | 59.3% | 352 | 40.7% |  |
| Other children with the same mother |  |  |  |  |  |
| No | 629 | 60.7% | 407 | 39.3% | <0.001 |
| Yes | 704 | 47.6% | 776 | 52.4% |  |
| Integration into the healthcare system |  |  |  |  |  |
| Rarely or never in contact with a physician | 458 | 53.8% | 394 | 46.2% | 0.609 |
| Has a primary care physician or chronic disease follow-up | 875 | 52.7% | 786 | 47.3% |  |

<sup>1</sup> When the sum of the category headcounts does not correspond to the total headcount, there is missing data (see table 1).

#### Men’s characteristics associated with their attendance

After adjustment for other men’s characteristrics, father attendance remained strongly associated with the couple’s expecting a first child (odds Ratio [OR] 2.00 95% confidence interval [95% CI] 1·67-2·40*, p<0*·*001*) and with the father’s being born abroad (Subsaharan Africa OR 1·83 95% CI 1·45-2·31, Asia OR 1·82 95% IC 1·26-2·63, figure 2, graph a).

Men aged 30 and over remained more receptive to the offer than younger men. The same U-shaped curve was observed for educational level: men with little or no schooling and men with higher education were more likely to seek advice than men who had studied up to secondary school. Fathers’ age was associated with their compliance in the global study population (figure 2, graph b).

#### Immigrated men’s characteristics associated with their attendance

In multivariate analysis, immigrants from Subsaharan Africa and Asia attended more than those from Europe or North America (OR 1.96 [1.33-2.88] and OR 2.06 [1.28-3.34], respectively), while paternal age was no longer associated with the acceptance rate. Immigrant men involved in a recent relationship came more often than those in a longer relationship (gifure 2, graph d).

After adjustment for other precariousness criteria including country of origin, healthcare coverage, length of stay and employment, undocumented fathers remained more likely to attend than fathers with residence permit (OR 2·41, 95% CI 1·80-3·22 in the complete multivariate model and OR 1·93, 95% CI 1·34-2·77 in the final model, additional file 3).

**Figure 2:**
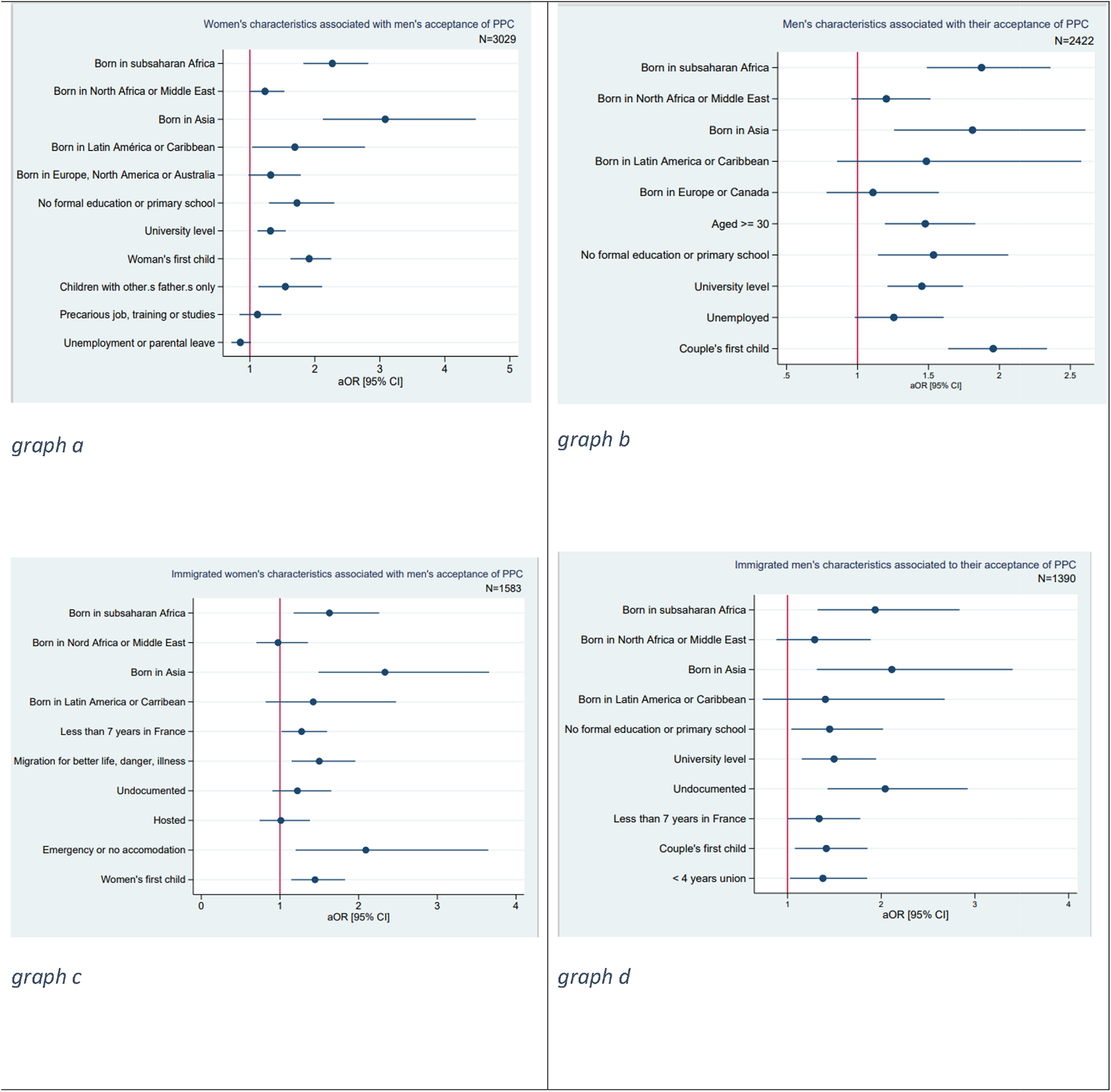
Women’s and men’s characteristics associated with men’s acceptance of Father’s Prenatal Prevention Consultation (FPPC), Montreuil (France), 2021-2022

## Discussion

Our main findings were a high level of acceptance by men (44% of those contacted, 53% of those with whom contact had been made), though lower than the women’s predictions (78% thought their partner would come). Acceptance was higher among immigrants, especially the most precarious ones.

### A high participation rate, based on a proactive approach

Taking as denominator the men targeted, participation rate was 44%. This is higher than the 36% participation rate in Seine Saint Denis for the breast cancer screening campaign, which has been launched in 2004 and would thus be expected to be more easly accepted (26). An other long-running offer, colorectal cancer screening, has an even lower acceptance rate in our seeting, especially in men (global 27%, men 25%)(27).

It has been shown that cancer screening was lower in socially disadvantaged and immigrants populations (28) (29). Thus, interventions based on proactive contact by navigators to guide vulnerable patients through the health care system and address barrier that may prevent them from getting the prevention and care they need have been built in the USA (30). Navigators call patients on the phone, explain the screening, schedule appointments, assess lack of transportation, financial and insurance barriers: the effectiveness of such programs in increasing screening rates has been demonstrated for breast, cervical and colorectal cancers (31). Earlier, proactive interventions also proved their efficiency in promoting HIV testing in future fathers, in different Subsaharan African settings: randomized controlled trials showed higher acceptance rates when fathers received an invitation letter (12), sometimes with an appointment already booked (32), and when caregivers came to the couples’ home (33).

Our proactive approach of calling men, adapting to their needs by offering them different appointment times and locations and rescheduling missed consultations probably played a major role in the high level of acceptance. However, we have always sought consent from pregnant women before contacting fathers, in order not to disempower women (34). Although the association did not remain significant after adjustment, the partners of women who reported a first HIV test during the current pregnancy were more likely to come than those of women who were unaware of having been tested, and than those of women who had been tested previously. This finding supports the promotion of couple-oriented screening. However, using the symbolic occasion of childbirth to incite fathers to care for their own health would probably not be sufficient without this active outreach, as long as the social norm on both parent’s duties remains unchanged (35). Even if the gendered distribution of responsabilities makes women the guardians of children’s health, prenatal care has not always been a standard: in France, pregnant women’s visits were initially a condition for the granting of family benefits (36).

### Immigrants were the most receptive to the offer, especially those with social vulnerabilities

Immigrant men took up the offer better than French-born men, particularly those from Subsaharan Africa and Asia. This differential acceptance persisted after adjustment to men’s social vulnerabilities. Therefore, the most precarious immigrants are the ones who came to the FPPC the most.

Yet the reduced flexibility of working hours for fathers in precarious employment has been identified as a potential obstacle to their attendance at classic antenatal visits (37), the availability issue was less frequently raised by socially vulnerable men than by well established ones (unshown data:reasons given by eligible fathers when refusing the offer) .

One possible explanation for immigrants’ greater attendance is their need to be helped in opening up health insurance rights: 7% of men had no health insurance coverage when surveyed, although French system theoretically covers nearly everyone. Undocumented immigrants living in France for at least 3 months can apply for State Medical Assistance (SMA), a separate system that covers the entire cost of care, but they are often unaware of their entitlement to SMA or how to obtain it (38). Another explanation could be greater uncovered healthcare needs in immigrants (39).

Surprisingly, while lack of health insurance coverage was associated with the use of consultations, having a medical follow-up was not. Number of French-born men with no social vulnerability did not have any medical follow up and did not attend FPPC. We may hypothesize that the reason why well-integrated men do not take up the offer is that they have the social skills to access the healthcare system, should a health problem occur. This hypothesis is supported by the finding of inequitable access to the healthcare system for immigrants in various European countries, including France (40).

### Strenghts and limitations

While most of the research published in the field has emphasised the importance of involving men in prenatal care in order to improve the health of the mother and the development of the child, this is, to our knowledge, the first structured healthcare intervention built in the perspective of improving fathers’ own health.

This survey has several limitations: research team’s outreach efforts towards the fathers, free and easy access to a multidisciplinary team, may not be replicable in routine clinical practice. Moreover, monocentric design limits the generalizability of the results. Given our context, marked by social insecurity and immigration, we reached men who may not have the same needs as the general population of fathers. We can, however, project that in areas where poverty prevails, the most disadvantaged men would seize a routine prenatal consultation as a point of entry into the health system.

## Conclusion

Although a prenatal consultation with biological tests is theoretically possible in France, it is not yet implemented. This study has shown that such consultation is well accepted when, for the first time, it is actually organized and offered. The men who make most use of it are immigrants with social vulnerabilities, and those who have been in France for less time. This consultation therefore appears as an opportunity for men facing administrative obstacles, or who do not have the social codes of the country they live in, to integrate into the healthcare system. Scaling up this intervention might thus reduce social inequalities in health. Consideration now needs to be given on how to make it available and free of charge in current practice.

## Data Availability

All data produced in the present study are available upon reasonable request to the authors

https://classic.clinicaltrials.gov/ct2/show/NCT05085717

## List of abbreviations

MQ: Maternal Questionnaire
SMA: State Medical Assistance (Aide Médicale de l’Etat in French)
FPPC: Father’s Prenatal Prevention Consultation
CI: Confidence Interval OR: Odd Ratio

## Declarations

### Ethics approval and consent to participate

The French Data Protection Authority (CNIL, registration number 921135) and the French Personnal Data Protection Comittee (Comite de Protection des Personnes, registation number 21.01.19.44753 amended by 21.05022.944753 decision, allowing and increased number of inclusions and additional data collection) gave full ethical approval. Pregnant women and fathers-to-be included in the study gave their consent after being fully informed

### Consent for publication

Participants’ consent to publication is not required, as no identifying information is provided. All co-authors have reviewed and approved the submitted manuscript.

### Availability of data and materials

The datasets generated during PARTAGE study are not publicly available, but are available from the corresponding author on reasonable request.

### Authors’ contributions

Conceptualization: PP, ADL, GJ, AG, TP

Data curation: PP, TP, RH, AG, GJ, CT

Formal analysis: PP, ADL

Funding Acquisition: PP, PEM

Investigation: VAM, CT, GJ, AG

Methodology: PP, ADL

Project administration: PP, GJ, AG, PEM, BR

Software: TP

Supervision: ADL

Writing-original draft: PP, ADL

## Aknowledgments

The authors would like to thank all the people who participated in the study. The PARTAGE Study Group included, in addition to the authors: Anne-Laurence Doho, Patricia Obergfell, Djamila Gherbi, Emilie Daumergue, Anne Simon, Miguel de Sousa Mendes, Naima Osmani, Sandrine Dekens, Oumar Sissoko (ARCAT Association), Virginie Supervie, France Lert, Stéphanie Demarest, Ngone Diop.

We thank Christophe Michon, Nathalie Lydié, Joanna Orne-Gliemann, Laurent Mandelbrot, Corinne Taeron, Nicolas Derche, Gwenaelle Morvan, Bernadette Rwegera, Mélanie Jaunay, Ruth Foundje Notemi, Elisa Wardzala, Caroline Regnier, Paul Chalvin, Priscillia Ribouchon, Neima Sghiouar, Abdelkrim Imechket, Pauline Aubry, Francis Bouvier, David Benhammou, Andrainolo Ravalihasy, Karna Coulibaly, Guy Nielsen, the Montreuil Municipality, the Seine Saint Denis Departmental Council.

